# Associations between physical and mental health and the utilization of ambulatory and emergency healthcare among asylum-seekers and refugees. Results from a cross-sectional survey in Berlin, Germany

**DOI:** 10.1101/2022.06.01.22275809

**Authors:** Nora Gottlieb, Martin Siegel

## Abstract

**Background:** Asylum-seekers and refugees (ASR) exhibit high prevalence rates of chronic and mental illness, but low utilization of ambulatory specialist healthcare. Forgoing timely healthcare when facing formal and informal access barriers may direct ASR toward emergency care. This paper addresses the interrelations of physical and mental health and the utilization of ambulatory and emergency care, and explicitly addresses the associations between the different types of care.

**Methods:** A structural equation model was applied to a sample of n=136 ASR living in accommodation centers in Berlin, Germany. Patterns of emergency care utilization (outcome) and physical and mental ambulatory care utilization (endogenous predictors) were estimated, while controlling for age, sex, chronic conditions, bodily pain, depression, anxiety and length of stay in Germany (exogenous predictors) and self-rated health (endogenous predictor).

**Results:** Significant associations were observed between ambulatory care utilization and poor self-rated health (0.207, 95%-CI: 0.05; 0.364), chronic illness (0.096, 95%-CI: 0.017; 0.175) and bodily pain (0.019, 95%-CI: 0.002; 0.036); between mental healthcare utilization and anxiety (0.202, 95%-CI: 0.051; 0.352); and between emergency care utilization and poor self-rated health (0.621, 95%-CI: 0.059; 1.183), chronic illness (0.287, 95%-CI: 0.012; 0.563), mental healthcare utilization (0.842, 95%-CI: 0.148; 1.535) and anxiety (0.790, 95%-CI: 0.141; 1.438). We found no associations between ambulatory care utilization and emergency care utilization.

**Conclusions:** Our study generates mixed results concerning the associations between healthcare needs and ambulatory and emergency care utilization among ASR. We found no evidence that low utilization of ambulatory healthcare contributes to emergency care utilization among ASR; neither did we find any evidence that obtaining ambulatory treatment obviates the need to seek emergency care. Our results indicate that higher physical healthcare needs as well as anxiety are associated with more utilization of both ambulatory healthcare and emergency healthcare; whereas healthcare needs related to depression tend to remain unmet. Improving health services’ accessibility and responsiveness, including the expansion of support services, outreach, and the coverage of medical interpretation, may enable ASR to better meet their healthcare needs.

**Key Messages:** *Implications for policy makers:*

- We examined if low utilization of ambulatory healthcare among asylum-seekers and refugees (ASR) contributes to potentially avoidable and resource-intensive emergency room visits among this group.
- We did not find evidence that low ambulatory care utilization determines high emergency care utilization among ASR; neither did we find evidence that getting ambulatory treatment obviates the need to seek emergency care.
- Our study results show that some ASR patients use both ambulatory and emergency care, either moving back and forth between the two types of care (which suggests that neither one meets their need) or seeking either type of care “randomly” (which indicates problems navigating the health system). ASR with depression tend to not get any care for this problem.
- Our findings signal the need to improve accessibility and responsiveness of health services, including understandable health information, help with navigating the health system, low-threshold and outreach services, medical interpretation, and sensitization of administrative and medical health staff.
- Enabling ASR and other diverse groups to get specialized healthcare for their physical and mental health problems will contribute to better health system outcomes, including better health and less health inequalities, greater satisfaction among patients and staff, and more efficient healthcare provision, i.e. less preventable costs and burdens for the health system.

*Implications for public:* Refugees often have difficulties in getting the healthcare they need. We tested if this makes them use more emergency care. This would be problematic for patients and healthcare providers; for example, because emergency services are already strained and costly. Indeed, our study suggests that refugees with anxiety go back and forth between ambulatory and emergency care, maybe because neither service resolves their problems. ASR with a chronic disease also use both ambulatory and emergency care. It is good if people with more health needs use more healthcare; it is even better, though, if we ensure they get specialized services for their particular problems. Refugees with depression tend to not get any help. More outreach, support with accessing the right healthcare provider, interpretation services and intercultural training for staff will help refugees get better care; and it will help healthcare providers offer treatment for refugees and other minorities effectively and efficiently.

## Background

Despite substantial health needs, asylum-seekers and refugees (ASR) in Germany exhibit lower utilization of ambulatory health services than statutorily insured persons as regards ambulatory specialist healthcare and ambulatory mental healthcare (1–8). In light of comparatively high incidence rates of emergency room visits and avoidable hospitalizations among ASR in Germany (1,2,4,5,7), this raises concern: Formal and informal access barriers may make ASR forgo timely treatment in the ambulatory sector and instead use other, potentially inadequate and more costly emergency health services (4,9,10). Incomplete information about ASRs’ medical history and healthcare needs among health service providers may exacerbate such patters, leading to inadequate, insufficient or misguided provision of healthcare (11,12). Discrepancies between healthcare needs and the treatment provided, in turn, are liable to frustrate care-givers and patients and increase costs (13,14). In the long run, inadequate treatment of physical and mental health conditions compromises the wellbeing, quality of life, social relations, and integration of ASR in the host country, and entails economic losses, for example through reduced productivity or resources spent on informal care (9,15).

This paper investigates the potential associations of physical and mental healthcare needs with ambulatory and emergency healthcare utilization among ASR. It employs a Structural Equation Model (SEM) to data generated by a cross-sectional survey of ASR in Berlin. To our knowledge, this is the first paper that addresses the role of ambulatory physical and mental healthcare utilization for the utilization of emergency care by ASR by explicitly modeling potential interdependencies between the utilization of the different types of care.

### Healthcare needs of asylum-seekers and refugees

ASR are at particular risk for physical and mental ill health because of structural factors before, during and after displacement (16–18). Infectious diseases and, to a certain extent, mental trauma have been the primary foci of health system responses in transit and destination countries. However, healthcare provision for ASR may thus tend to overlook further important healthcare needs such as specialized services for chronic non-communicable diseases, sexual and reproductive health, or dental health (18,19). Especially (absent health services for) chronic non-communicable diseases have been described as an emerging challenge for ASR in the context of recent forced migratory movements such as from Syria and Ukraine (20–22).

Owing to a lack of systematic monitoring and routine data on ASRs’ health in Germany, comprehensive information on healthcare needs in this population is unavailable (23). Empirical studies indicate that communicable diseases such as acute respiratory infections are a frequent reason for seeking healthcare among ASR; prevalence rates of communicable and parasitic diseases are comparable to the general German population (4,24). As regards the prevalence of chronic non-communicable diseases, study results differ: Some authors report relatively low rates of chronic illness among ASR, while stating that findings may be due to underreporting/-diagnosing (6,24). Others show similar or higher prevalence rates for chronic diseases as compared to the general population in Germany (5,25); Bauhoff and Göpffarth (4), for example, found prevalence rates of 20%, 48% and 62% respectively for nutritional anemia, diabetes and hypertension (as compared to 9%, 50% and 62% among the general population). High rates of unspecific symptoms and conditions that are often related to psychological distress, such as chronic back pains and headaches, have been linked to potential somatization (4,6,24,26). This is consistent with high prevalence rates of mental distress and illness – up to 77% for post-traumatic stress disorder (27) and around 45% for depression and anxiety (4–6) respectively – that were found among ASR in Germany. Studies consistently report overall low subjective health (5,6).

### Accessibility of healthcare for asylum-seekers and refugees

Treatments to improve ASRs’ physical and mental health outcomes exist (28). Community-based provision of the respective services has proven particularly effective (29,30). To realize the potential of these services, it is crucial to ensure accessibility and continuity of care through universal health coverage, low-threshold supply of services, case management and health navigation services, availability of interpreters and bilingual staff, as well as close intersectoral collaboration (31,32). However, healthcare provision for ASR in host countries is often restricted to acute care. Utilization of healthcare for chronic and mental health conditions is frequently hampered by eligibility restrictions and requires out-of-pocket payments. Missing interpretation services, low health literacy among ASR, and related difficulties with navigating complex healthcare and referral systems create further barriers to healthcare utilization (19). With regard to mental health, fear of stigmatization and discrepant expectations and beliefs concerning mental health and healthcare may further impede utilization (9,33).

In Germany, ASR only become eligible for a full (i.e., equivalent to statutory health insurance) scope of health services after a waiting period of 18 months from their time of arrival in the country (34). Until then, their health coverage is limited to the treatment of acute and life-threatening conditions. Treatment of chronic diseases and mental health conditions can be covered by the social welfare office after an individual case review (34); but ASR may experience substantial hurdles during this procedure. For instance, 49% of ASRs’ applications for psychotherapy coverage were rejected after the individual case review, as compared to a rejection rate of 6% among statutorily insured persons (35). Across Germany, ASR may face different modes of healthcare provision, and thus different levels of health service accessibility, because the actual organization of healthcare provision for ASR is at the discretion of the local authorities (36). In Berlin, ASR obtain an electronic health insurance card upon arrival, which, in theory, should allow for access to healthcare providers in a similar fashion as statutory health insurance (13).

In addition to formal barriers, ASR may experience various informal barriers, including low availability and acceptability of health services. In this context, considerable communication barriers and sometimes negative attitudes from the part of administrative and medical staff have been reported (37–39). They have been linked to insufficient coverage of medical interpretation services, a lack of intercultural training for medical staff, and general deficits in the accommodation of cultural and social diversity in healthcare provision in Germany (40,41). In mental healthcare, specifically, a shortage of healthcare providers poses problems. Although specialized “Psychosocial Centers” (in German: Psychosoziale Zentren, PSZs) offer mental healthcare services for ASR, in addition to psychotherapists and psychiatrists, existing capacities cannot meet the demand (41). In 2019, the PSZs reported waiting times of up to two years for psychotherapy and a 40% rejection rate due to a lack of capacities (35). In addition, stigma toward mental illness and healthcare among some ASR groups creates obstacles to needs-based mental healthcare utilization (39,42).

## Methods

### Study design and sampling

This study uses a cross-sectional survey on the health needs and healthcare utilization among ASR. The target population were ASR living in shared accommodation centers in Berlin, who were of legal age, and who were able to complete the questionnaire in one of the nine languages provided. Administering the survey in accommodation centers offers the opportunity to obtain a representative sample of the heterogeneous asylum-seeking population in Germany, because German law obliges ASR to reside in shared accommodation centers for the first 18 months of their stay or until obtaining permanent residency status. Given the scarcity of affordable housing, many ASR remain in shared accommodation centers beyond the designated period.

A clustered randomized sampling approach was applied to include a representative distribution of accommodation centers in the study: Using a complete list of Berlin’s accommodation centers, the facilities were divided into three categories, according to their capacity (small facilities with less than 250 persons, medium facilities with 250-500 persons, and large facilities with over 500 persons). The distribution of ASR across the different categories was calculated (19% in small centers, 59% in medium centers, 22% in large centers) and proportional numbers of accommodation centers were drawn from each category to achieve a similar distribution in the study sample.

Accommodation centers were contacted via email and telephone. If no contact could be established or participation in the survey was rejected, a new accommodation center from the same category was drawn. Within each participating accommodation center, the research team endeavored to obtain the highest possible number of respondents.

### The questionnaire

This study is based on a shortened version of a questionnaire developed in the RESPOND project (“Improving regional health system responses to the challenges of migration through tailored interventions for asylum-seekers and refugees”)((5)). The version used here comprised 55 items including I) health status, II) healthcare utilization, and III) sociodemographic information.

I. Health status included self-assessed health status (measured on a Likert scale from 1=very good to 5=very poor), chronic illness *(“Do you have any longstanding illnesses or health problems?”*), screening items for depression (PHQ-2) and anxiety (GAD2), and a six-point scale for bodily pain.
II. Utilization of healthcare within the preceding 12 months (yes/no) was surveyed for general practitioners, specialist practitioners, psychotherapists, and psychiatric care providers, as well as for emergency care. The number of emergency room visits in the last 12 months was assessed.
III. Sociodemographic information included gender (female/male/diverse), age, legal status (asylum application pending/asylum application concluded with refugee status/subsidiary protection/humanitarian protection/rejection), length of stay in Germany in months, and the highest level of formal education accomplished.

The questionnaire and all related information material were available in nine languages (Albanian, Arabic, English, Farsi, French, German, Russian, Serbian, and Turkish).

### Ethical approval

Ethical clearance was obtained from the Ethics Committee of the Charité University Clinic (EA4/111/18).

### Data collection

Data were collected between June 2018 and December 2019. The research team announced the study to staff and residents of the participating accommodation centers via posters and written information in different languages, and, if possible, in person, for example during in-house plenary sessions. Depending on local conditions, the questionnaire was administered in two different ways: In some accommodation centers, the research team went from door to door and invited the residents to participate in the survey. In other centers, the team positioned itself in a public area of the accommodation center and invited passers-by to participate. In either case, symbolic giveaways were offered irrespective of study participation. Study information was provided in writing and verbatim with the help of recorded explanations in different languages, if necessary. The research team tried to avoid any pressures to participate in the study, for example by emphasizing that the decision whether or not to participate would neither effect the asylum procedures nor involve any other personal risks or benefits. The questionnaire was handed out on paper for independent completion, together with a stamped envelope. Filled-out questionnaires were returned in three different ways: 90% were handed over in person, 5% were deposited in a closed box in the accommodation center, and the remaining 5% were sent by mail.

### Variables

The variables used for the empirical analysis included binary variables for the presence of a chronic disease, symptoms of depression and symptoms of anxiety, for utilization of ambulatory physical healthcare in the preceding 12 months, utilization of ambulatory mental healthcare in the preceding 12 months, and for less-than-good (i.e., very poor, poor or fair) self-assessed health. Bodily pain was included on a five-point Lickert scale. The number of emergency care visits in the preceding 12 months is the main outcome of this study.

### Statistical Analysis

A Structural Equation Model (SEM) was employed to investigate the potential interrelations of physical and mental health, of ambulatory physical and mental healthcare utilization, and of emergency healthcare utilization. The SEM allows a simultaneous estimation of the associations of physical and mental health needs with the utilization of different types of health services. This is considered necessary here because the different services are neither mutually exclusive nor independent from one another. A set of regressions with one-dimensional outcomes may therefore lead to biased results, when neglecting the expected correlations of ambulatory physical and mental healthcare utilization with the utilization of emergency healthcare. The SEM further allows modelling the associations between health and healthcare utilization as both direct and indirect effects. This is done here to reveal how having accessed ambulatory physical and mental healthcare in the preceding 12 months may shape the pattern of emergency healthcare utilization.

The estimation model shown in Figure 1 is built as a set of direct and indirect associations between health and healthcare utilization. Poor self-assessed physical health is included as an endogenous predictor of ambulatory physical healthcare and emergency healthcare utilization, with chronic illness, bodily pain and age serving as exogenous predictors of self-rated physical health. Symptoms of depression and anxiety are considered as exogenous predictors of ambulatory mental healthcare and emergency care utilization. Direct paths are included from ambulatory physical and mental healthcare utilization to emergency care utilization to assess whether ambulatory care utilization is associated with emergency care utilization. Indirect paths are modelled from fair or poor health and from the presence of chronic illness to ambulatory physical and emergency care utilization, as well as from the presence of depressive symptoms and the presence of anxiety to emergency care utilization. Gender and less than 18 months of stay in Germany are included with direct effects on all types of healthcare utilization, to control for potential effects of the scope of health entitlements and familiarity with the German healthcare system on utilization patterns. All estimations were performed in Stata 15.1 using the SEM command.

**Figure 1:**
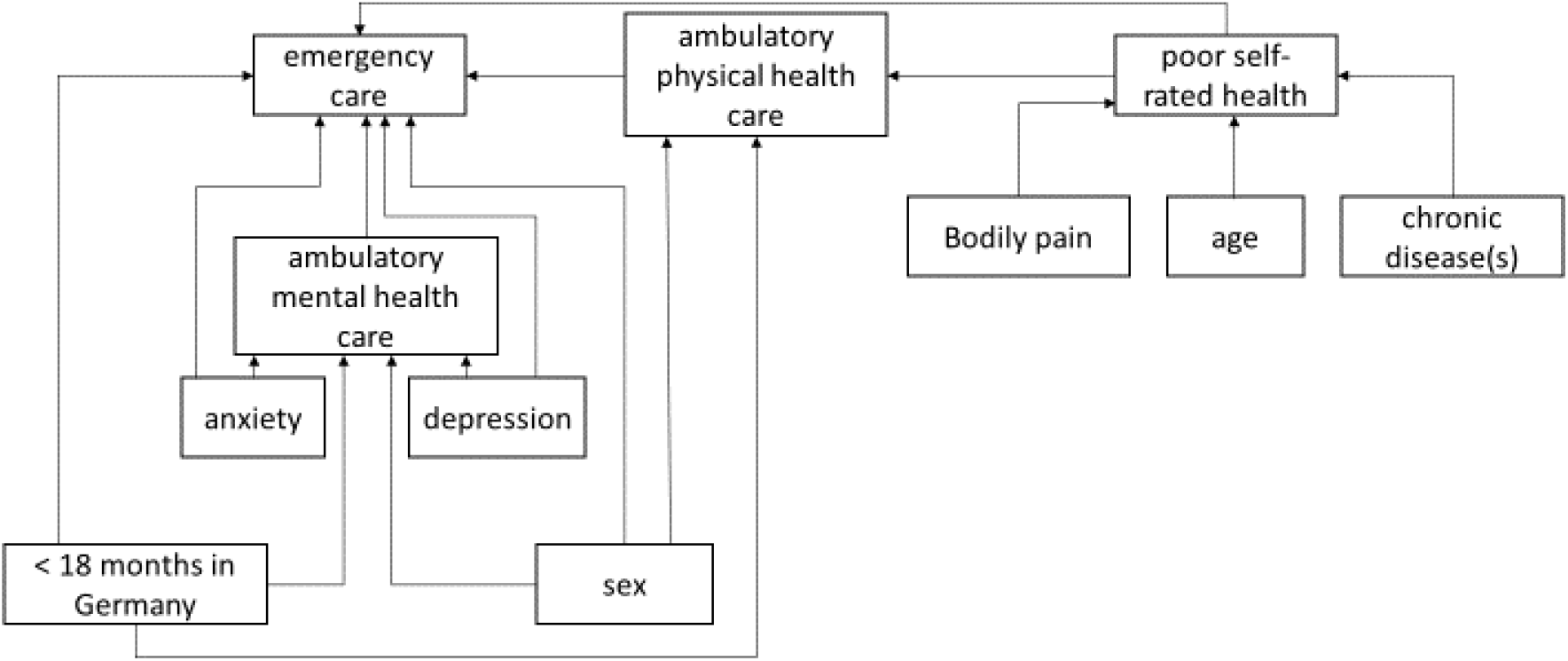
Illustration of our full estimation model, built of direct and indirect associations between physical and mental health, ambulatory physical and mental healthcare utilization, and emergency healthcare utilization among asylum-seekers and refugees

## Results

### Descriptive results

Twenty-two out of 74 accommodation centers for ASR in Berlin participated in this study. At the time, 6,399 ASR were residing in shared accommodation centers in Berlin. Among them, an estimated 3,839 (60%) met the inclusion criteria for this study. Out of 811 persons who could be approached, 327 filled out and returned the questionnaire, which corresponds to a cooperation rate of 39% and a response rate of 8%. Upon data cleansing, a total N=309 observations remained. Not all respondents completed all questionnaire items; our sample for a complete case analysis therefore comprises 136 observations.

Respondents were on average 34 years old, 37% described their gender as female and 63% as male. The average time since arrival in Germany was 39 months. 25% of the respondents arrived in Germany less than 18 month ago; their scope of healthcare entitlements therefore underlies restrictions. Almost half (46%) of the respondents described their physical health as less than good; 38% report at least one chronic illness. Symptoms of at least one mental illness were reported by approx. 50% of the respondents; where 39% reported only depressive symptoms, 38% symptoms of anxiety, and 27% symptoms of both.

More than half of the estimation sample had had at least one ambulatory care visit within the preceding 12 months. Only 21% of respondents report at least one mental healthcare visit in the previous year. One third of the respondents used emergency healthcare at least once within the preceding 12 months, the average yearly number of emergency healthcare utilizations among this group was 2.4. In the overall estimate sample, the average number of reported emergency care utilizations was 0.79. The descriptive results of the analysis are summarized in Table 1.

**Table 1:** Description of the estimation sample

|  |  | min | max |
| --- | --- | --- | --- |
| Mean age (in years) | 33.90 | 18 | 68 |
| Sex (female) | 36.76% |  |  |
| Less than good health | 45.59% |  |  |
| At least one chronic disease | 37.50% |  |  |
| Mean score of bodily pain scale (1=low, 5=high) | 2.44 | 1 | 6 |
| Symptoms of depression | 38.97% |  |  |
| Symptoms of anxiety | 38.24% |  |  |
| Stay in Germany (in months) | 39.18 | 2 | 159 |
| Fraction of respondents with stay < 18 months in Germany | 25.00% |  |  |
| Fraction of respondents who used ambulatory care in the preceding 12 months | 55.88% |  |  |
| Fraction of respondents who used psychiatric care or psychotherapy in the preceding 12 months | 21.32% |  |  |
| Average number of emergency care visits in the preceding 12 months | 0.79 | 0 | 10 |

N=136

### Estimation results

The likelihood-ratio-test indicates no significant difference between the observed and the estimated covariance structure of the data, suggesting that the estimated associations between health and healthcare utilization reflect the underlying interrelations. In addition, the Non-Normal (Tucker-Lewis) Fit Index is above 0.95 and the comparative fit index is above 0.9, both indicating a very good model fit. The root mean squared error (RMSEA) of 0.034 is below the threshold of 0.05, which, too, suggests a very good model fit. The model fit indexes are presented in Table 2.

**Table 2.** Goodness of fit indexes

| goodness of fit index | value | threshold |
| --- | --- | --- |
| Likelihood-ratio-test (model vs. observed) | 18.536 (p=0.293) | p>0.05 |
| RMSEA | 0.034 | < 0.05 |
| Comparative Fit Index | 0.982 | >0.95 |
| Tucker-Lewis (non-normal) Fit index | 0.961 | >0.9 |
| AIC | 3305.124 | n/a |
| BIC | 3380.853 | n/a |

Figure 2 illustrates the estimation model after all insignificant paths were removed. The estimates for the direct, indirect and total associations between the variables are presented in Table 3. The structural equation for self-rated health suggests that bodily pain and the presence of a chronic disease are associated with poor self-assessed health. The estimated coefficients of chronic disease and pain are both significantly positive, indicating that the risk to rate one’s health as fair, poor or very poor is higher if a respondent reports at least one chronic disease or bodily pain.

**Table 3:**
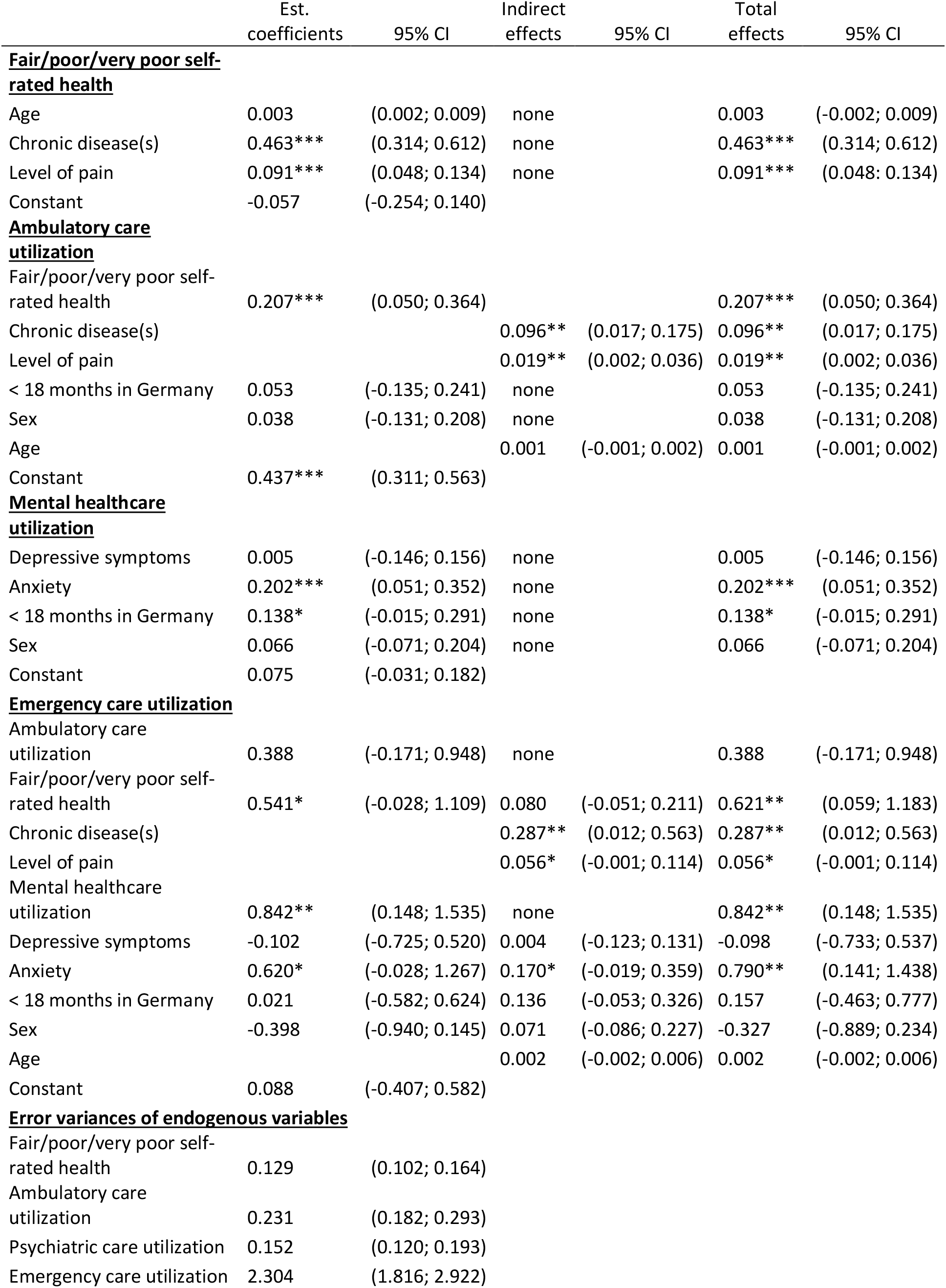
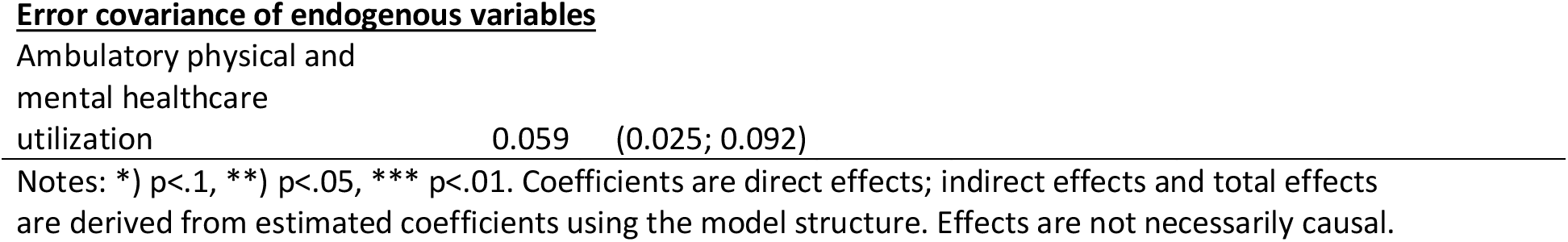
Estimation results of the direct and indirect associations between physical and mental health, ambulatory and mental healthcare utilization, and emergency healthcare utilization among asylum-seekers and refugees

|  | Est.<br>coefficients | 95% CI | Indirect<br>effects | 95% CI | Total<br>effects | 95% CI |
| --- | --- | --- | --- | --- | --- | --- |
| **<u>Fair/poor/very poor self-rated health</u>** |  |  |  |  |  |  |
| Age | 0.003 | (0.002; 0.009) | none |  | 0.003 | (-0.002; 0.009) |
| Chronic disease(s) | 0.463*** | (0.314; 0.612) | none |  | 0.463*** | (0.314; 0.612) |
| Level of pain | 0.091*** | (0.048; 0.134) | none |  | 0.091*** | (0.048; 0.134) |
| Constant | -0.057 | (-0.254; 0.140) |  |  |  |  |
| **<u>Ambulatory care utilization</u>** |  |  |  |  |  |  |
| Fair/poor/very poor self-rated health | 0.207*** | (0.050; 0.364) |  |  | 0.207*** | (0.050; 0.364) |
| Chronic disease(s) |  |  | 0.096** | (0.017; 0.175) | 0.096** | (0.017; 0.175) |
| Level of pain |  |  | 0.019** | (0.002; 0.036) | 0.019** | (0.002; 0.036) |
| < 18 months in Germany | 0.053 | (-0.135; 0.241) | none |  | 0.053 | (-0.135; 0.241) |
| Sex | 0.038 | (-0.131; 0.208) | none |  | 0.038 | (-0.131; 0.208) |
| Age |  |  | 0.001 | (-0.001; 0.002) | 0.001 | (-0.001; 0.002) |
| Constant | 0.437*** | (0.311; 0.563) |  |  |  |  |
| **<u>Mental healthcare utilization</u>** |  |  |  |  |  |  |
| Depressive symptoms | 0.005 | (-0.146; 0.156) | none |  | 0.005 | (-0.146; 0.156) |
| Anxiety | 0.202*** | (0.051; 0.352) | none |  | 0.202*** | (0.051; 0.352) |
| < 18 months in Germany | 0.138* | (-0.015; 0.291) | none |  | 0.138* | (-0.015; 0.291) |
| Sex | 0.066 | (-0.071; 0.204) | none |  | 0.066 | (-0.071; 0.204) |
| Constant | 0.075 | (-0.031; 0.182) |  |  |  |  |
| **<u>Emergency care utilization</u>** |  |  |  |  |  |  |
| Ambulatory care utilization | 0.388 | (-0.171; 0.948) | none |  | 0.388 | (-0.171; 0.948) |
| Fair/poor/very poor self-rated health | 0.541* | (-0.028; 1.109) | 0.080 | (-0.051; 0.211) | 0.621** | (0.059; 1.183) |
| Chronic disease(s) |  |  | 0.287** | (0.012; 0.563) | 0.287** | (0.012; 0.563) |
| Level of pain |  |  | 0.056* | (-0.001; 0.114) | 0.056* | (-0.001; 0.114) |
| Mental healthcare utilization | 0.842** | (0.148; 1.535) | none |  | 0.842** | (0.148; 1.535) |
| Depressive symptoms | -0.102 | (-0.725; 0.520) | 0.004 | (-0.123; 0.131) | -0.098 | (-0.733; 0.537) |
| Anxiety | 0.620* | (-0.028; 1.267) | 0.170* | (-0.019; 0.359) | 0.790** | (0.141; 1.438) |
| < 18 months in Germany | 0.021 | (-0.582; 0.624) | 0.136 | (-0.053; 0.326) | 0.157 | (-0.463; 0.777) |
| Sex | -0.398 | (-0.940; 0.145) | 0.071 | (-0.086; 0.227) | -0.327 | (-0.889; 0.234) |
| Age |  |  | 0.002 | (-0.002; 0.006) | 0.002 | (-0.002; 0.006) |
| Constant | 0.088 | (-0.407; 0.582) |  |  |  |  |
| **<u>Error variances of endogenous variables</u>** |  |  |  |  |  |  |
| Fair/poor/very poor self-rated health | 0.129 | (0.102; 0.164) |  |  |  |  |
| Ambulatory care utilization | 0.231 | (0.182; 0.293) |  |  |  |  |
| Psychiatric care utilization | 0.152 | (0.120; 0.193) |  |  |  |  |
| Emergency care utilization | 2.304 | (1.816; 2.922) |  |  |  |  |

**Error covariance of endogenous variables**
|  |  |  |
| --- | --- | --- |
| Ambulatory physical and<br>mental healthcare<br>utilization | 0.059 | (0.025; 0.092) |

Notes: \*) $p < .1$ , \*\*) $p < .05$ , \*\*\*) $p < .01$ . Coefficients are direct effects; indirect effects and total effects are derived from estimated coefficients using the model structure. Effects are not necessarily causal.

**Figure 2:**
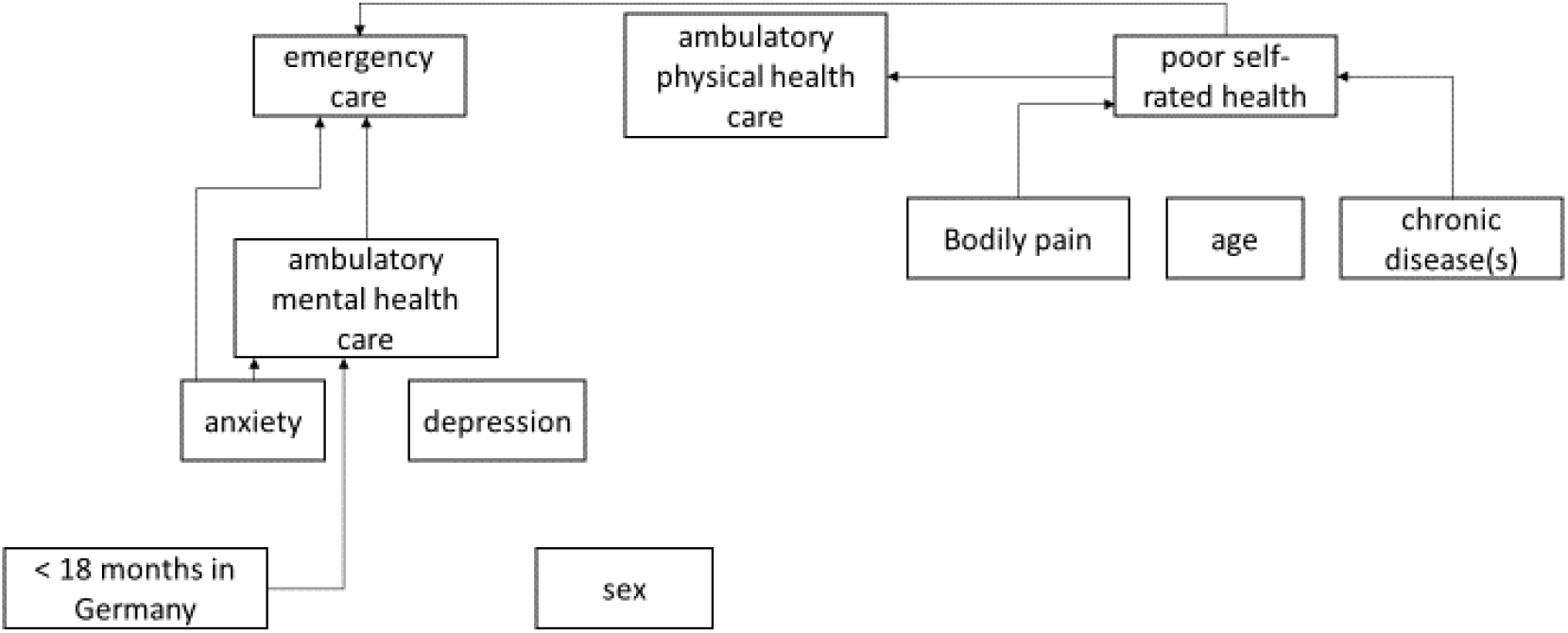
Illustration of our estimation model, showing only significant associations between physical and mental health, ambulatory physical and mental healthcare utilization, and emergency healthcare utilization among asylum-seekers and refugees

Only fair, poor or very poor self-rated health was directly and significantly associated with ambulatory physical healthcare utilization. A statistically significant indirect association was found for chronic disease and bodily pain with ambulatory care utilization. The likelihood of ambulatory mental healthcare utilization is significantly higher at a 1% level among ASR who screened positive for anxiety, and it is significantly higher for those who have been in Germany for less than 18 months, but only at a 10% level.

Emergency care utilization yielded no significant associations with ambulatory physical healthcare utilization, but we found a significant association of emergency care utilization with mental healthcare utilization. Further variables that were associated with a higher frequency of emergency care utilization are chronic illness, bodily pain, and anxiety. The presence of chronic illness is indirectly associated with a higher frequency of emergency care utilizations through its association with poor self-rated health at a 5% significance level. Likewise, bodily pain also has a significant indirect association with the utilization of emergency care services at the 10% level.

Having symptoms of anxiety shows a strong and significant positive association with the utilization of both ambulatory mental healthcare and emergency care. In contrast, symptoms of depression have no significant association with the utilization of neither ambulatory mental healthcare nor emergency care. Gender and length of stay exhibit no significant associations with emergency care utilization.

## Discussion

The aim of this study was to disentangle the interrelations between physical and mental healthcare needs, the utilization of ambulatory physical and mental healthcare services, and the utilization of emergency care by ASR. In light of various formal and informal barriers to healthcare faced by this population, it has been suggested that foregone ambulatory healthcare visits may contribute to high emergency care utilization (4,9,18). Our study was the first to explicitly account for the non-exclusiveness and potential interdependence of the two types of care among ASR in Germany. The results indicate that ambulatory mental healthcare utilization, alongside poor self-rated health, chronic illness, and anxiety, is significantly associated with increased emergency care utilization. The utilization of ambulatory physical healthcare and the presence of depressive symptoms were not associated with emergency care utilization.

With regards to the question whether low utilization of ambulatory healthcare drives emergency care use among ASR, our study yields mixed results, which may reflect differences in healthcare seeking for different health problems. Overall, however, it is important to note that we found no significant negative associations between the utilization of any (physical or mental) ambulatory healthcare and emergency care utilization, as we would expect either a) if the foregoing of necessary visits to an ambulatory healthcare provider would eventually result in (avoidable) emergency room visits; or b) if health problems were resolved in ambulatory care, thus obviating the need for a visit to the emergency room.

As regards mental health, our results confirm the discrepancy between ASRs’ high burden of illness and their relatively low utilization of ambulatory mental health services, which has been reported in the international literature and which suggests that ASR with mental health needs may generally be at risk of remaining underserved (33). Our study indicates that this may be especially the case regarding depression: The finding that depressive symptoms are not associated with the utilization of any type of healthcare service points toward unmet need for this condition. Anxiety disorder, on the contrary, is associated with an increased likelihood of using both ambulatory mental healthcare and emergency care. The positive association between the utilization of both ambulatory mental healthcare and emergency care may indicate that ASR patients tend to go back and forth between the two types of service, with neither one meeting their need. Our study thus supports other authors’ claims that mismatches between ASRs’ mental health needs and the healthcare provided for them contributes to high utilization rates of other health services, incl. emergency care (43,44).

As regards physical health, we found higher utilization of both ambulatory and emergency healthcare among ASR with higher physical healthcare needs. This could indicate a desirable outcome from the perspective of vertical health equity. However, it has been pointed out that high utilization rates of emergency services, in combination with frequent emergency room visits for non-severe conditions and during non-social hours, may rather reflect either inaccessibility of regular healthcare, or failures of ambulatory health services to meet ASRs’ needs (4,18). It has been argued, for example, that excessive painkiller prescription for ASR patients signal a tendency to relief symptoms instead of attending thoroughly to patients to sustainably cure the problem (3,45). Our findings lend only limited support for these claims (as we would then expect to find a negative association between ambulatory and emergency care utilization in case of inaccessibility, or a positive association in case of a mismatch of healthcare needs and provision). Another explanation may be that difficulties in navigating a bureaucratic, complex and fragmented healthcare system, in combination with a lack of understandable health information (6,37), make ASR patients rather “randomly” seek either ambulatory healthcare, emergency healthcare, or both types of care to resolve physical ailments. Overall, our results underline that, to develop a sufficiently nuanced understanding of health needs and healthcare utilization patterns among ASR populations that can inform health policy and planning, future research needs to include ASRs’ perspectives (18,19).

Similar to previous studies (46), we found that respondents who had arrived in Germany within 18 months before the survey were more likely to report utilization of mental healthcare. ASR who had arrived more recently are arguably less familiar with the German healthcare system. However, the coverage of medical interpretation services during the first 18 months of stay in Germany may facilitate access to mental healthcare. This entitlement expires after 18 months (6,34). Another explanation for our finding is that the centralized accommodation of ASR during the first 18 months – despite its manifold disadvantages – may facilitate the provision of social support, including encouragement to seek mental healthcare and help with navigating the healthcare system. Recent research on ASRs’ perspectives on mental healthcare provision in Germany suggests that support services via official (e.g., social workers) and unofficial channels (e.g., NGO workers, volunteers, and peers) play a key role in facilitating access to mental healthcare services (39). ASR who move out of centralized accommodation centers may be at risk of discontinuing treatment once they drop out of these support networks. This indicates the need for institutionalized and expanded support services, which can facilitate needs-based healthcare seeking and continuity of care in a comprehensive and sustainable fashion (12). The recent displacement of large numbers of persons from Ukraine and their decentralized settlement across the member states of the European Union renders this need all the more urgent. Ultimately, such change has the potential to render the health system more accessible and responsive for diverse population groups, including, albeit not restricted to ASR, and thus contribute to better health system outcomes for all.

This study has three major limitations. First, the sample size is comparatively small. This limits the generalizability of the results and leads to low statistical power, which, in turn, may result in insignificant results even if a systematic association exists. However, the sample size is sufficiently large for the employed SEM; and we consider the final estimation sample of 136 complete cases to be a good success in data collection, given the manifold challenges in conducting empirical research with ASR (47,48). Second, the questionnaire asked for utilization of ambulatory healthcare, ambulatory mental healthcare, and emergency care over a period of 12 months, which may involve recall bias. However, the potential of errors arising from this are expected to be rather small. Finally, the study was restricted to the German city-state of Berlin. Availability of interpretation services and healthcare providers with compatible language skills, as well as regulations of healthcare provision within the first 18 months of stay vary considerably across Germany, thus limiting the generalizability of our findings. However, given the similarity of research findings from different contexts, the results of this study may provide some general insights into healthcare needs and utilization patterns among ASR.

## Conclusions

We found no clear evidence for low ambulatory healthcare utilization acting as a determinant of high emergency care utilization among ASR. However, our study supports previous claims concerning the German healthcare system’s deficits in terms of accessibility and responsiveness of service provision for ASR populations. These deficits, in turn, may induce undesirable healthcare seeking patterns such as patients going back and forth between ambulatory and emergency care, or an undirected (“random”) utilization of the two types of services. ASR with depressive symptoms are at particular risk of remaining underserved. To develop a nuanced understanding of the needs and healthcare seeking of ASR with chronic and mental illness, better data and further research, especially studies that include ASRs’ perspectives, are required. Improving accessibility and responsiveness of healthcare services, including the expansion of institutionalized support, outreach, and coverage of interpretation services beyond ASRs’ first 18 months in the country, may be key to improving both health outcomes among ASR and health system outcomes, including greater efficiency of healthcare provision for a diverse society.

## Disclaimers

### Source(s) of support/funding

The research leading to these results has received funding from the People Programme (Marie Curie Actions) of the European Union’s Seventh Framework Programme (FP7/2007-2013) under REA grant agreement no. 600209 (TU Berlin/IPODI). Furthermore, we acknowledge support for the publication costs by the Open Access Publication Fund of Bielefeld University and the Deutsche Forschungsgemeinschaft (DFG).

### Disclosure of relationships and activities

The authors have no conflicts of interest to declare.

### Ethical issues

Ethics Committee of the Charité University Clinic Berlin (IRB no. EA4/111/18)

## Data Availability

All data produced in the present study are available upon reasonable request to the corresponding author.

## Acknowledgements

The authors would like to thank all study participants for their valuable time, and the accommodation centers’ staff and management, the Berlin State Office for Refugee Affairs and the Berlin Senate Administration for their support of this study. We thank the RESPOND team for making the questionnaire available and for further assistance, and to all students of the Berlin School of Public Health who contributed to the survey administration.

## Authors’ contributions

obtaining funding: NG; conception and planning: NG; data collection: NG; statistical analysis: MS; analysis and interpretation: NG, MS; drafting of manuscript: NG, MS; review and approval of manuscript: NG, MS

